# Parametrization of Worldwide Covid-19 data for multiple variants: How is the SAR-Cov2 virus evolving?

**DOI:** 10.1101/2024.04.09.24305557

**Authors:** Dietrich Foerster, Sayali Bhatkar, Gyan Bhanot

## Abstract

We mapped the 2020-2023 daily Covid-19 case data from the World Health Organization (WHO) to the original SIR model of Karmack and McKendrick for multiple pandemic recurrences due to the evolution of the virus to different variants in forty countries worldwide. The aim of the study was to determine how the SIR parameters are changing as the virus evolved into variants. Each peak in cases was analyzed separately for each country and the parameters: r_eff_ (pandemic R-parameter), L_eff_ (average number of days an individual is infective) and α (the rate of infection for contacts between the set of susceptible persons and the set of infected persons) were computed. Each peak was mapped to circulating variants for each country and the SIR parameters (r_eff_, L_eff_, α) were averaged over each variant using their values in peaks where 70% of the variant sequences identified belonged to a single variant. This analysis showed that on average, compared to the original Wuhan variant (α = 0.2), the parameter α has increased to α = 0.5 for the Omicron variants. The value of r_eff_ has decreased from around 3.8 to 2.0 and L_eff_ has decreased from 15 days to 10 days. This is as would be expected of a virus that is coming to equilibrium by evolving to increase its infectivity while reducing the effects of infections on the host.

## Introduction and Method

Extensive data on the spread of the SARS-Cov-2/Covid-19 epidemic from 2020 to the present for most countries are available from the World Health Organization (WHO) [1]. Here we consider three years of data, from 3/1/2020 to 28/2/2023, for a subset of countries where the peaks in daily identified infections from various variants separate into well-defined peaks. Various mathematical models of epidemics have been described in the literature, with the SIR model, published nearly a century ago [2], being the earliest and simplest of such models (see [3,4] for reviews).

Before one can determine whether a mathematical model describes the available WHO data, one encounters the difficulty that the WHO data describes only reported (symptomatic and/or tested) infections, while the true number of infections remains unknown. Extended SIR models have been proposed to resolve this difficulty [5,6,7,8,9]. In this study, we parametrize the ratio of reported infections to the total number of infections as a constant for a given peak but variable for peaks from different variants of the virus within each country. We also assume that waves of infections from variants can be treated as independent, with an SIR model describing each of them separately. With these two simple hypotheses, we show that the SIR model provides a qualitatively correct parametrization of most of the world’s SARS-Cov-2/Covid-19 data.

In the SIR model of epidemic infections [2], the number of infected I(t) and susceptible S(t) individuals in a population of N interacting individuals evolves as a function of time t (measured in days) according to the following coupled ordinary differential equations:

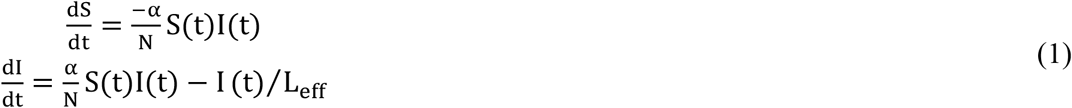

Here α is the rate of infection between an infected person in the I compartment and a susceptible person in the S compartment, and L_eff_ is the time an infected person remains infective. For each peak, we impose the initial conditions I(−∞) = 0, S(−∞) = N. We also assume that S(t), I(t) evolve sufficiently slowly for the derivatives 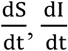to be meaningful. The number of parameters reduces to a single variable r_eff_ when these equations are rewritten in terms of the rescaled variables s = S/N, i = I/N and τ = t/L_eff_:

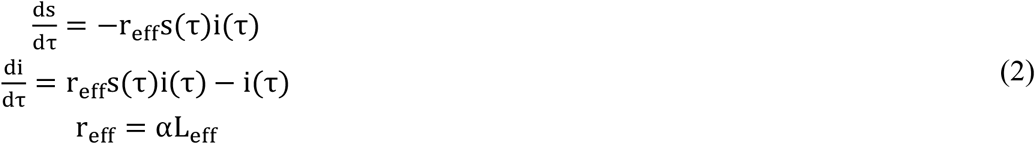

The initial conditions are now: i(−∞) = 0, s(−∞) = 1. From Eq. 2, it is easy to show that the number of infections i(τ) increases as exp(r_eff_− 1)τ for small τ, reaches a maximum at 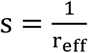 and decays as exp(−(1 − r_eff_s(∞))τ) for large τ.

### Reported symptomatic infections versus the total number of infections

As noted above, the WHO Covid-19 data reports only identified (symptomatic and/or tested) infections, whereas the SIR model in the form represented above describes all infections. To circumvent this difficulty, we assume that the infections reported by the WHO data represent, for a given peak and country, a fixed ratio of all infections. In other words, we represent the observed data i_observed_(t) in terms of model data i_model_(t, r_eff_, L_eff_) only up to a fixed multiplicative factor. Thus, we assume that the logarithms of the observed and model data differ by a constant c:

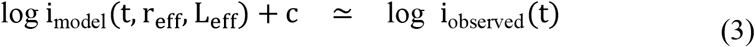

We will consider those parameters r_eff_, L_eff_ as “optimal” which minimize the average quadratic difference in the logarithms. Thus, we define the error:

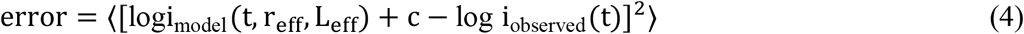

where ⟨ ⟩ indicates an average over the days of the epidemic wave under consideration. Since c is a function of r_eff_ and L_eff_, we minimize the error (Eq. 4) with respect to these two parameters.

### Decomposition of a country’s Covid-19 data into distinct peaks

The Covid-19 epidemic occurred in successive waves that are recognized as being due to distinct variants of the original virus (https://ourworldindata.org/grapher/covid-variants-area). An important source of noise comes from the data being collected and combined from different locations in different ways and reported at different times (days of the week) in different countries. Consequently, to identify the parameters for the various peaks, we had to first smooth the WHO data for I(t) by locally averaging them over a few days. We then used the maxima and minima for the smoothed data to decompose them into distinct peaks. Figure 1 shows the results of this smoothing procedure applied to the Covid-19 data of South Korea and Japan.

**Figure 1:**
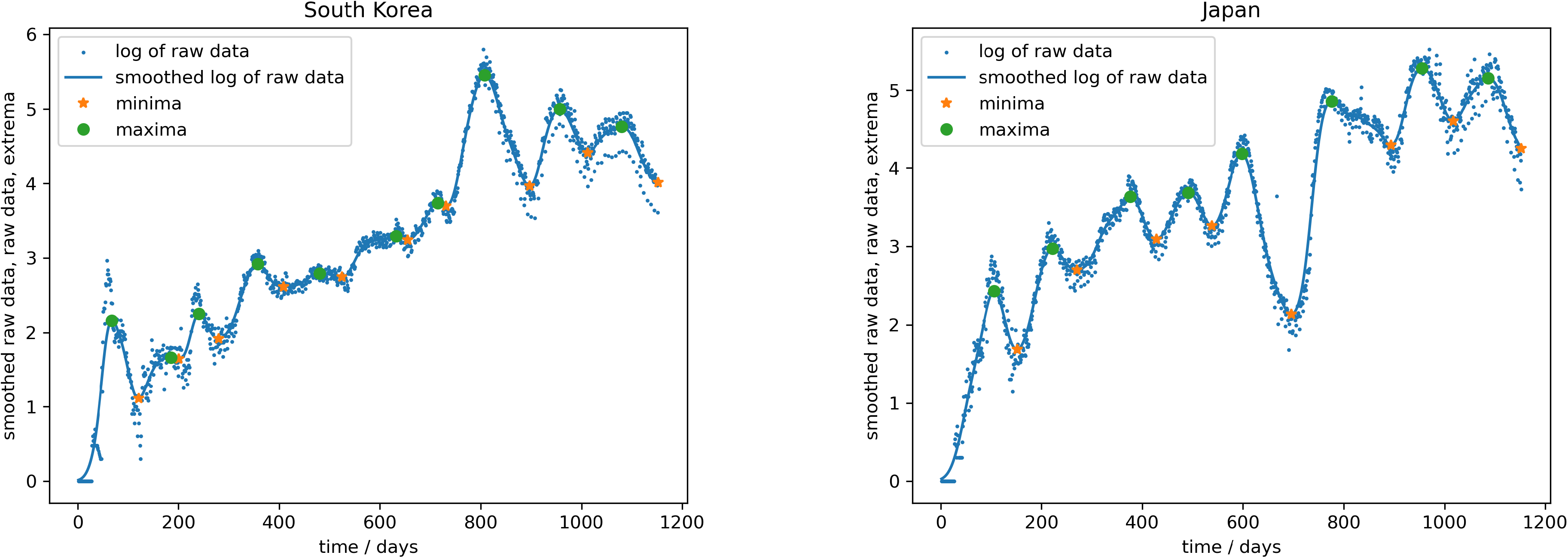
The figure shows the location of maxima and minima for two countries, South Korea and Japan. The minima locate the boundaries in time between which fits to the SIR model were made for each peak as described in the text.

### Measuring the parameters

We determined the optimal parameters γ_eff_ = 1/L_eff_, r_eff_ for each peak in the original raw data, using the maxima of the smoothed data as centers (t = 0) of the data. We do this by minimizing the quadratic error (Eq. 4) using the Powell minimization algorithm [10,11]. The results of this procedure for a number of countries are shown in Figure 2a,b,c. The complete results for all forty countries analyzed are in Supplementary Figures.

**Figure 2a-e:**
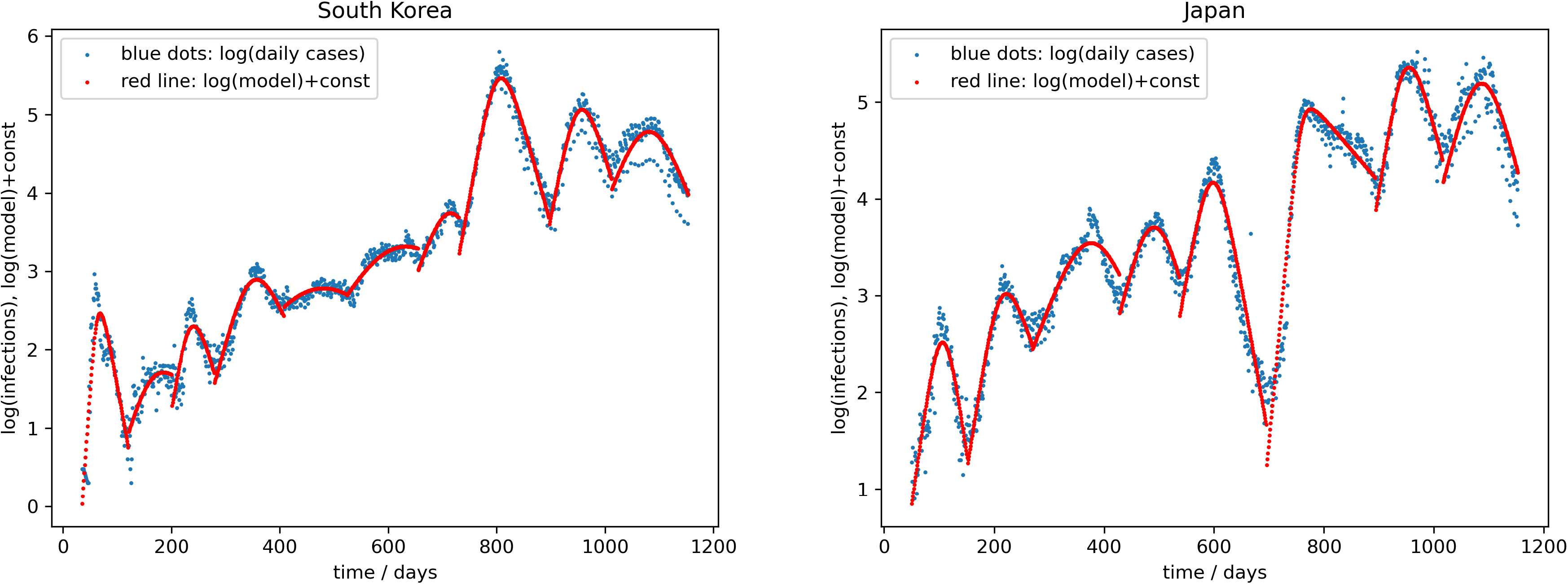
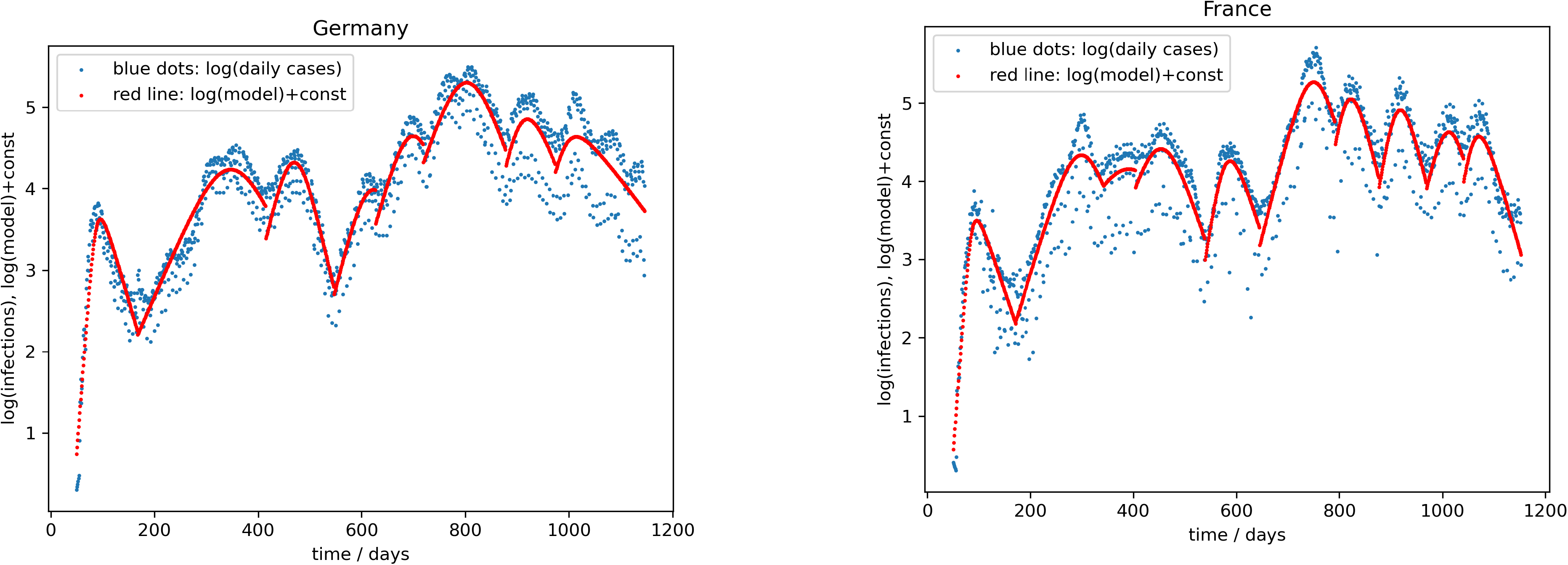
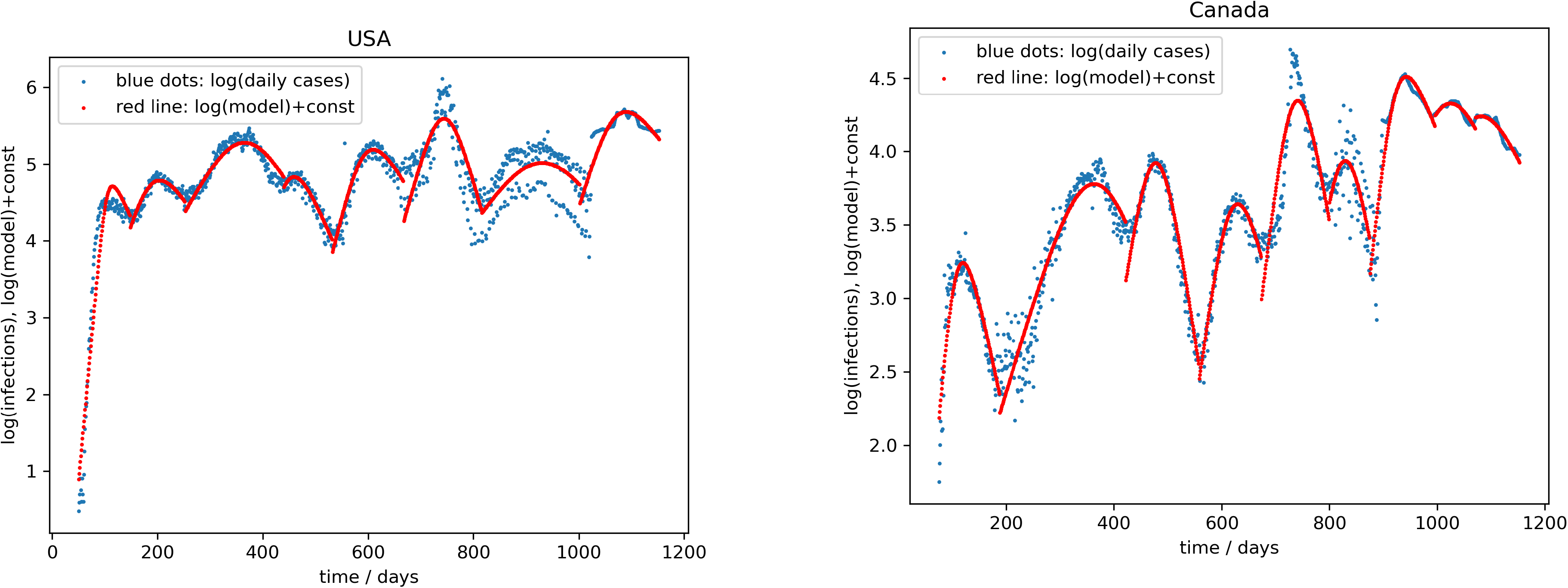
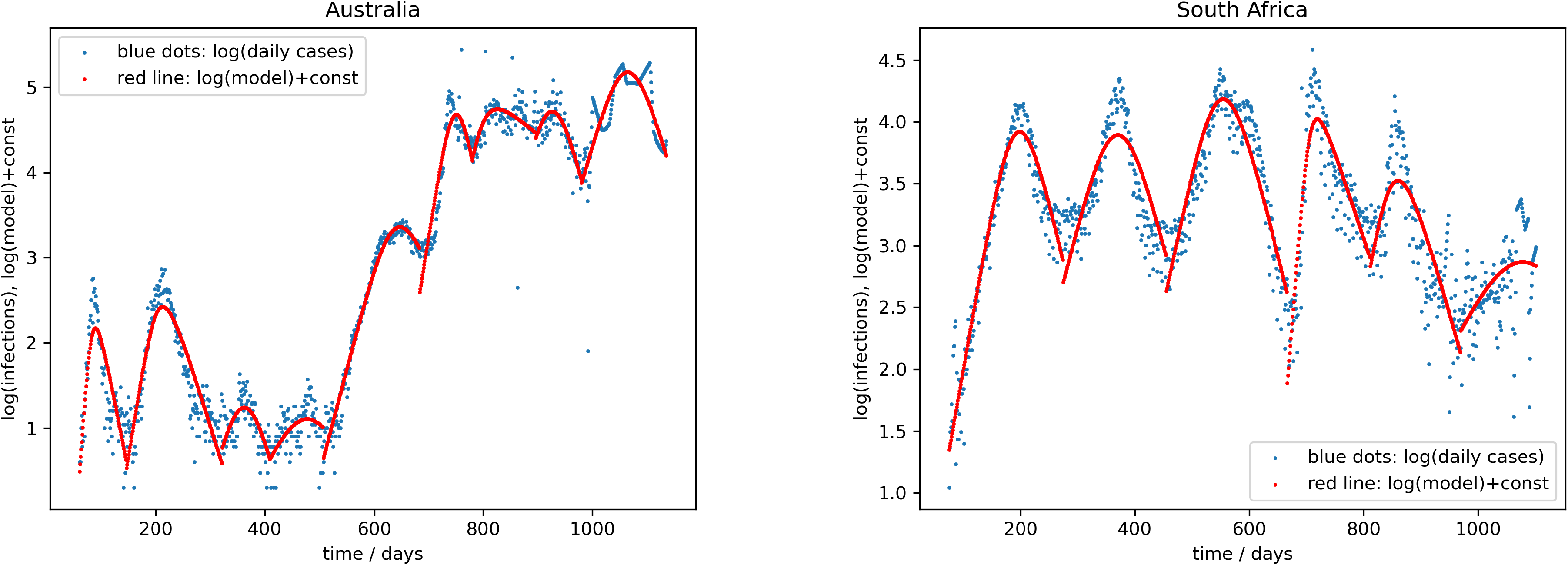
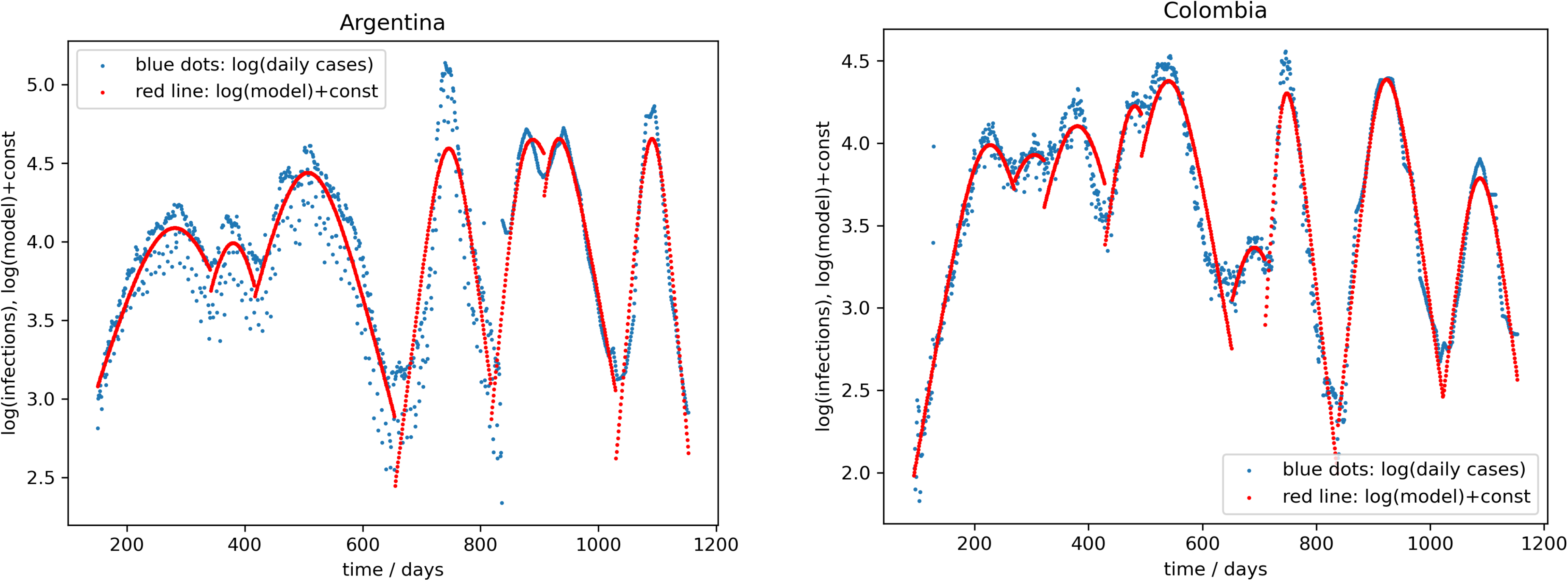
Shows the WHO data and the SIR model fits for ten countries, chosen from six continents.

To estimate the accuracy of our results, we added normally distributed noise to the averaged logarithm of the data:

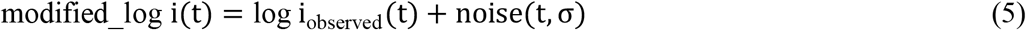

with σ taken as the mean deviation between the logarithm of the smoothed data and the logarithm of the raw data. The resulting dispersion of the parameters γ_eff_, r_eff_ provides an estimate of their accuracy. Because we optimized the parameters for the individual peaks, the transitions from one (computed) peak to the next were abrupt and show cusps or jumps in the derivative, while the observed WHO data are smooth because they do not distinguish between different virus variants.

### Details of Method used to solve the SIR equations

There are different ways to solve these equations. Our own procedure uses the known values of S, I at the peak as an anchoring point. In the equations we rescale variables as s(t) = S (t)/r_eff_ and i(t) = I (t)/r_eff_ to obtain

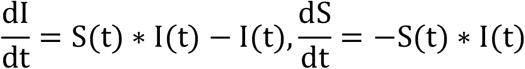

Now the dependence on r_eff_ is only via the initial condition S(−∞) = r_eff_. Using the above equations, it is now easy to find a quantity that remains unchanged under evolution in τ

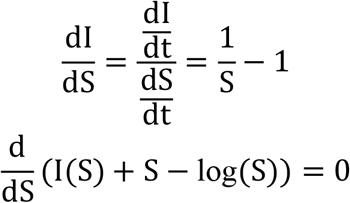

This implies that I(S) + S − log(S) remains unchanged under evolution in τ. Because 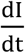 changes sign at S = 1 its maximum I_max_ must be at S = 1 and we obtain the well-known result:

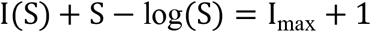

By specializing the last equation to t → −∞ and using S(−∞) = r_eff_ we find the actual value of I_max_

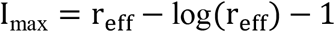

Using the point (S, I) = (1, I_max_) as a reference point, it is then easy to integrate the SIR equations in the positive and the negative τ directions using 4th order Runge-Kutta integration. Using a time step dτ = 0.01 we obtained results with an accuracy of better than 10^−10^ for the parameters and time ranges we used.

### Data used in the study

The subset of data for daily identified cases used in this study was obtained from WHO (https://covid19.who.int/WHO-COVID-19-global-data.csv) and is in Supplementary Table 1 and the data on variants identified for these countries as a function of time is in Supplementary Table 2 (from https://ourworldindata.org/grapher/covid-variants-area).

## Results

The SIR parameters r_eff_, L_eff_ and α from our analysis for forty countries are given in Supplementary Table 3. Supplementary Table 3 also shows the mapping of peaks in each country to one or more circulating variants at or near the peak, using data from Supplementary Table 2. SIR parameter values are given only for those peaks whose variants could be mapped.

Several peaks represented multiple variants, with varying fractions of measured variants (See Table 3). Peaks where a single dominant variant was circulating in over 70% of sequences from confirmed cases were identified from Supplementary Table 3 and are shown in Supplementary Table 4. We only retained those variants whose identity was confirmed in the data in Supplementary Table 2. For example, for India, the first peak was labeled as “non-who” so this data was not included in Supplementary Table 4. We identified peaks in all forty countries that had a single dominant variant whose measured fraction in Supplementary Table 2,3 was over 70%. The values of the parameters (r_eff_, L_eff_, α) were averaged for this variant over all countries for those peaks. This was done separately for each variant. Figures 3,4,5 show how these parameters changed on average from the original Wuhan variant to the variants that appeared chronologically worldwide: namely, Alpha, Beta, Gamma, Delta, and Omicron variants BA1, BA2, BA5 and BQ1. The variant Omicron BA4, which appeared almost at the same time as Omicron BA5 was subdominant in all peaks and countries we analyzed.

**Figure 3-5:**
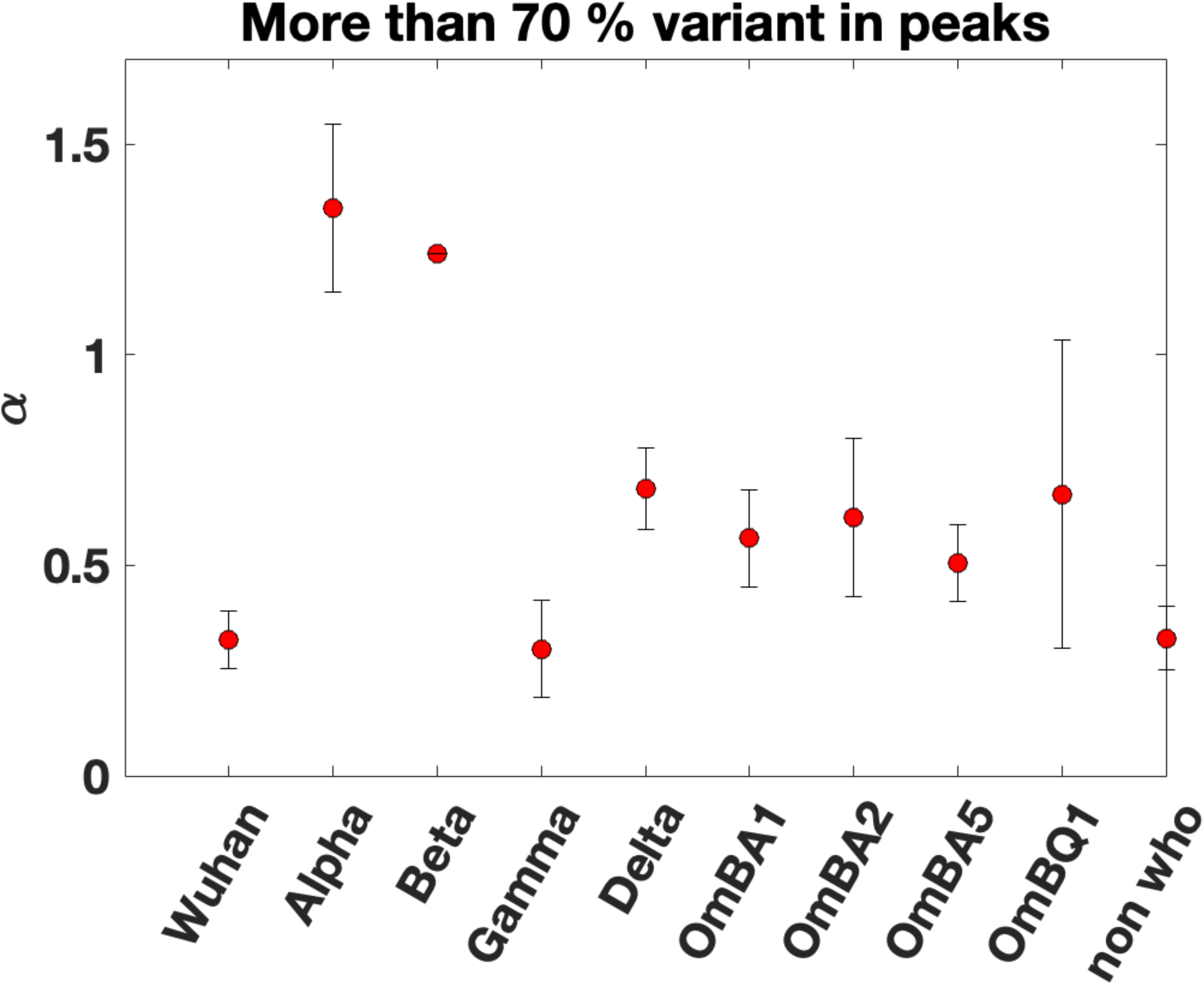
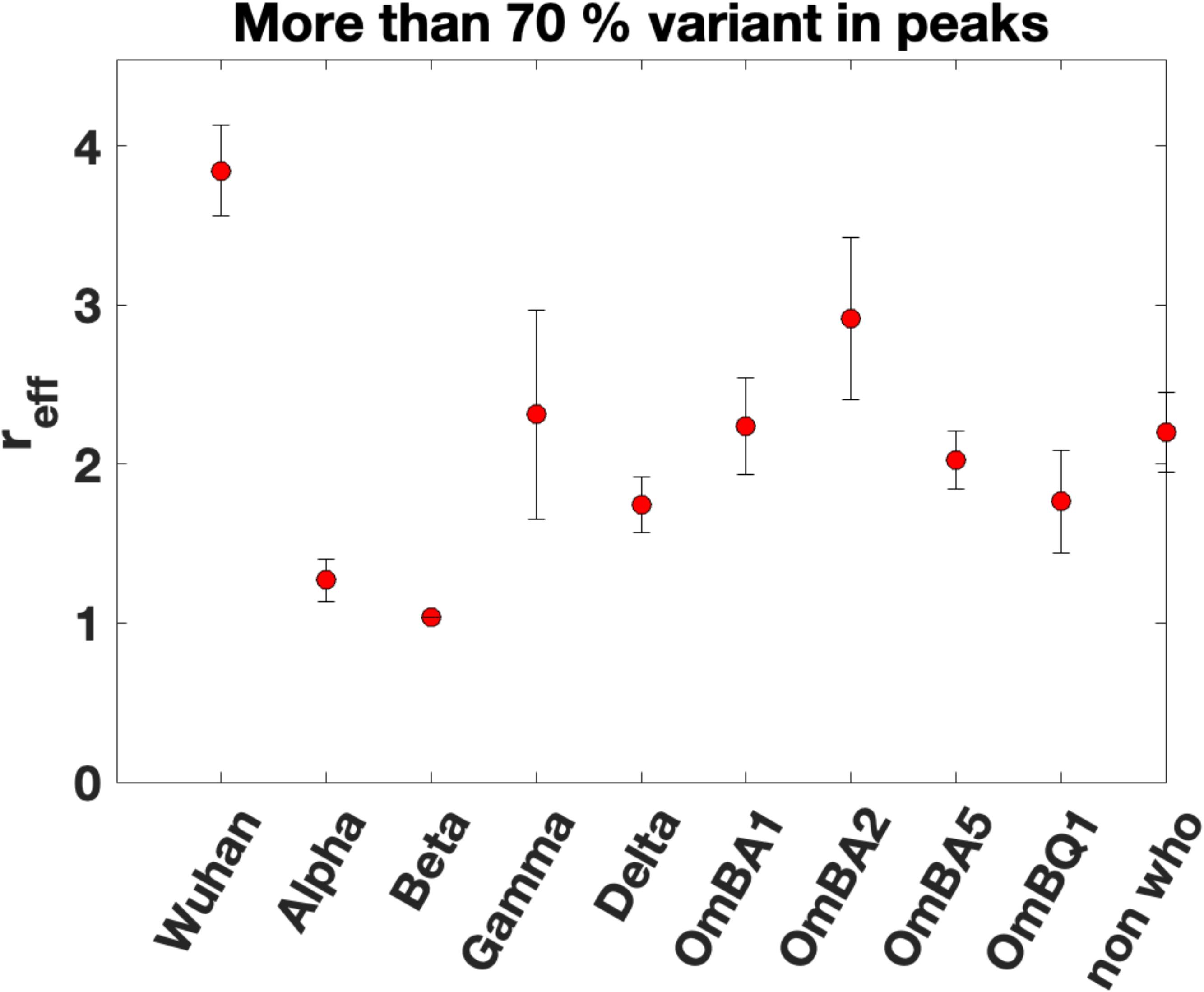
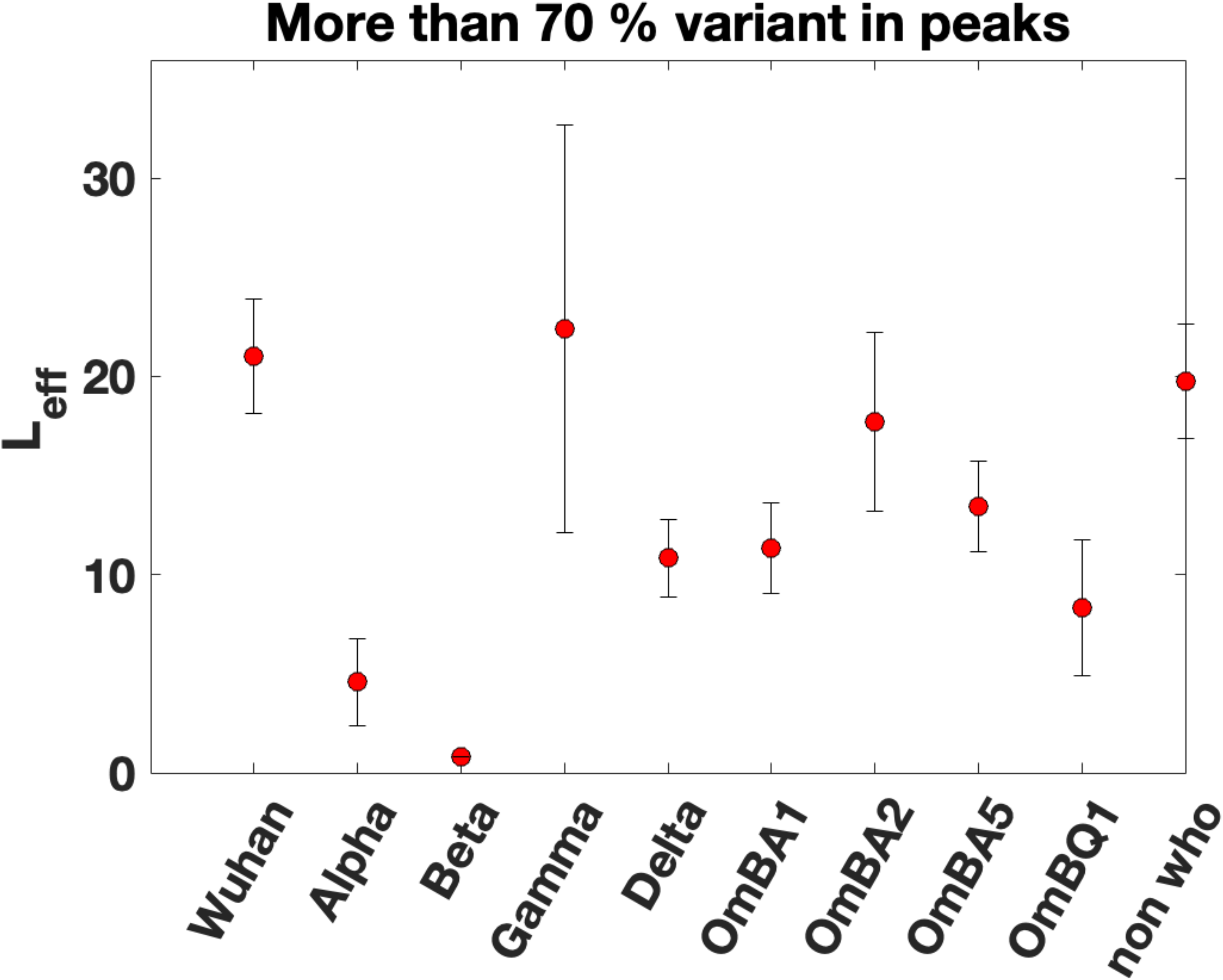
Show the worldwide variation of the SIR parameters α, r_eff_ and L_eff_ as a function of the Covid variant. For each variant, we averaged the SIR model parameters for peaks in daily cases where the variant represented more than 70% of the measured circulating virus. The x-axis approximately labels time as the variants are arranged in order of their appearance worldwide.

## Summary and Discussion

Several methods have been proposed to analyze multiple waves of pandemics from variants or strains. These methods try to identify mechanisms and their effects on various aspects of the pandemic. For example, in [12], the authors study five different mechanisms associated with multiple influenza epidemics. In contrast, [13] studies the effect of multi strain transmissions.

In this paper, we study the worldwide evolution of the Covid-19 pandemic as it developed into new variants. Our goal is to study the pandemic parameters as the virus evolved. To do this, we used worldwide WHO daily case data for Covid-19 (Supplementary Table 1) and analyzed it using the Kermack and McKendrick SIR model [2]. Each peak in cases was treated independently and the data mapped to the SIR Model yielded estimates of the SIR parameters r_eff_, L_eff_ and α. Some of the cut-points and fits to the data are shown in Figures 1 and 2 respectively. From the data on circulating variants measured worldwide (Supplementary Table 2), we chose forty countries which had the highest number of days with measurements of circulating variants. For these countries, the peaks were mapped to the data for circulating variants to identify the fraction of each variant type represented in the peak (Supplementary Table 3). For each variant, we identified peaks across all countries where a single dominant variant represented over 70% of cases (Supplementary Table 4). The SIR parameters for each such variant across all peaks and countries were averaged and are shown in Figures 3-5. The x axis in these figures approximately represents time because the variants listed are plotted in the order of their worldwide appearance.

Figures 3,4,5 show that on average, compared to the original Wuhan variant (α = 0.2), the parameter α dramatically increased in the Alpha and Beta variants but has since decreased to an asymptotic value close to α = 0.5 for the Omicron variants. The values of r_eff_ and L_eff_ have decreased from around 3.8 and 15 days respectively for the Wuhan variant to 2.0 and 10 days for the Omicron variants. This suggests that the SARS-Cov-2 virus is evolving to increase its infectivity (increase α) while reducing the values of r_eff_ and L_eff_.

Similar methods could be applied to other viral pandemics of the past and future.

## Data Availability

All data used is included in the supplementary material

## Author Contributions

All authors contributed equally to the development of the model and the analysis of the data. GB and DF wrote the paper. All authors have read and approved the manuscript, figures and supplementary materials.

## Acknowledgements

The authors thank world data collection agencies for providing public access to good data for our analysis.

## SUPPLEMENTARY FIGURE CAPTIONS

**Supplementary Figures:** Shows the WHO data and fits to the SIR model for all forty countries.

## SUPPLEMENTARY TABLE CAPTIONS

**Supplementary Table 1:** WHO data used in this study. The data was obtained from: https://covid19.who.int/WHO-COVID-19-global-data.csv.

**Supplementary Table 2:** Worldwide data on variants identified as a function of time. The data was obtained from https://ourworldindata.org/grapher/covid-variants-area.

**Supplementary Table 3:** SIR parameters from fits using methods described in the paper for all countries where each peak could be mapped to a circulating Covid variant using data from Supplementary Table 2.

**Supplementary Table 4:** SIR parameters for peaks where the measured circulating variant had at least 70% of a single variant.

